# AI-generated patient-friendly discharge summaries to empower patients

**DOI:** 10.1101/2025.07.04.25330804

**Authors:** Nils Reuter, Vincent-Noah von Lipinski, Jonathan Jeutner, Thorsten Schlomm, Martin Witzenrath, Leif Erik Sander, Matthias I Gröschel

## Abstract

**Background:** Patients often struggle to fully understand their discharge letters after inpatient hospital stays, which are often replete with domain-specific medical terminology. Large Language Models (LLMs) offer a promising solution by creating patient-friendly discharge summaries. However, direct evaluations of patient comprehension of such summaries have been limited.

**Methods:** This study explored whether AI-generated patient-friendly discharge summaries improve patients’ self-reported understanding of their medical condition. We provided patients at the night before discharge with AI-generated summaries of their discharge letters and recorded how this increased their understanding of their medical condition and procedures during their stay using an 11-item survey.

**Results:** Among hospitalized patients recruited at two departments in our tertiary care hospital (n=20), most (90%) reported better understanding after reading the AI-generated summary, including those who initially felt well-informed. Notably, 90% (18/20) found the summary more helpful compared to their discharge consultations, and older patients (above 69 years) showed particular interest in receiving such summaries for future hospital stays.

**Conclusion:** Our findings highlight the value of LLMs in improving patients’ comprehension of their medical condition. Larger studies are warranted to guide the implementation of patient-facing AI-generated content into healthcare.

## Background

It is well documented that patients recall little information from consultations or discharge interviews with physicians (1–3). During discharge from the hospital, patients receive written discharge letters replete with domain-specific medical terminology. Based on these letters, the patients are expected to attend follow-up consultations, navigate treatments, and manage new medications. In addition to the discharge letters, physicians routinely conduct discharge interviews, summarizing the treatment provided during the inpatient stay, addressing remaining questions and detailing the next steps. However, given limited physician time, these discharge interviews tend to be very brief and frequently leave patients unsatisfied and potentially confused (4).

Large Language Models (LLMs) can automatically generate patient-friendly discharge summaries from complex medical records and represent a promising tool to empower patients during discharge (5). Previous work showed that AI-generated summaries were easier to understand readability (6) and even outperformed physicians in summarizing medical content (7). However, these studies focused on readability scores or clinician ratings instead of recording patient-reported comprehension, empowerment, or satisfaction (8).

This study aimed to determine whether AI-generated patient-friendly discharge summaries can improve the patients’ understanding of their medical condition, including current diagnoses and care plans. We found that AI-generated summaries improved the patients’ self-reported health literacy, including patients with an already good understanding of their medical situation pre-intervention.

## Methods

### Large language models and prompt development

We used OpenAI’s GPT-4o (Version 2024-11-20 last trained October 2023) to generate our summaries as a widely adopted model that has shown strong clinical knowledge (9). We evaluated three different prompts to generate the summaries based on best practices in prompt engineering and using anonymized retrospective discharge summaries (10): First, a zero-shot prompt where we instructed the model without examples. Second, a one-shot prompt, providing the model with an example input and output. Third, a chain-of-thought prompt, suggesting the model steps to think through. Using each prompt, we generated six summaries from two separate discharge letters. Three physicians within at least their fourth year of training assessed the outputs using a single-blinded protocol where evaluators were unaware of which prompt produced which summary (Supplementary File S1). For the assessment, we used a 5-point Likert scale (1 = worst, 5 = best) across five domains: Relevance, Consistency, Simplification, Fluency, and Coherence.

### Patient survey and recruitment

We recruited 20 patients at two sites of Charité – Universitätsmedizin Berlin, a tertiary care center in Berlin, Germany, evenly split between internal medicine and surgery departments. Given this was an exploratory study, the cohort was a convenience sample that was not randomized or blinded.

The AI-generated summaries were obtained using the highest-performing prompting strategy based on the physician’s rating. Patients were given printed copies of the AI-generated summaries the evening before their hospital discharge. This timing ensured that patients had already received detailed information and discussed their condition with their physician (i.e., the discharge meeting). Patients were given an 11-item survey (Supplementary file S2), to self-evaluate their comprehension of their medical situation using a 6-point Likert scale (1 = Strongly disagree, 6 = Strongly agree). Items included the reason for the patients’ hospitalization, diagnostic procedures and their rationales, treatments received, and next steps regarding therapy, medication, and rehabilitation. The first six items of the survey evaluated the patients’ pre-intervention health literacy. After answering these items, patients were instructed to read the AI-generated summaries and to continue with the remaining 5 items of the survey, evaluating the post-intervention health literacy.

### Statistical analysis

All statistical analyses were conducted in R Version 4.4.3 (11). Data preprocessing, analysis, and visualization were performed using the tidyverse Version 2.0.0 (12). Responses from 6-point Likert-scale items were analyzed and visualized using the likert and ggstat packages (13,14).

Descriptive statistics were used to summarize participant demographics and survey responses. For Likert-scale items, means and standard deviations were calculated, and levels of agreement were expressed as percentages to capture overall response trends. Where appropriate, statistical tests were conducted to assess the significance of observed differences or associations.

## Results

### Prompt selection

All prompts yielded high scores among the three evaluating physicians, but the zero-shot prompt produced the most effective summaries (Figure 1A). While being ranked 0.5 points lower than the one-shot prompt in relevance, the zero-shot prompt was able to match other prompts in simplification and fluency and outperform them in coherence and consistency. We therefore continued to use the zero-shot prompt for our study.

**Figure 1.**
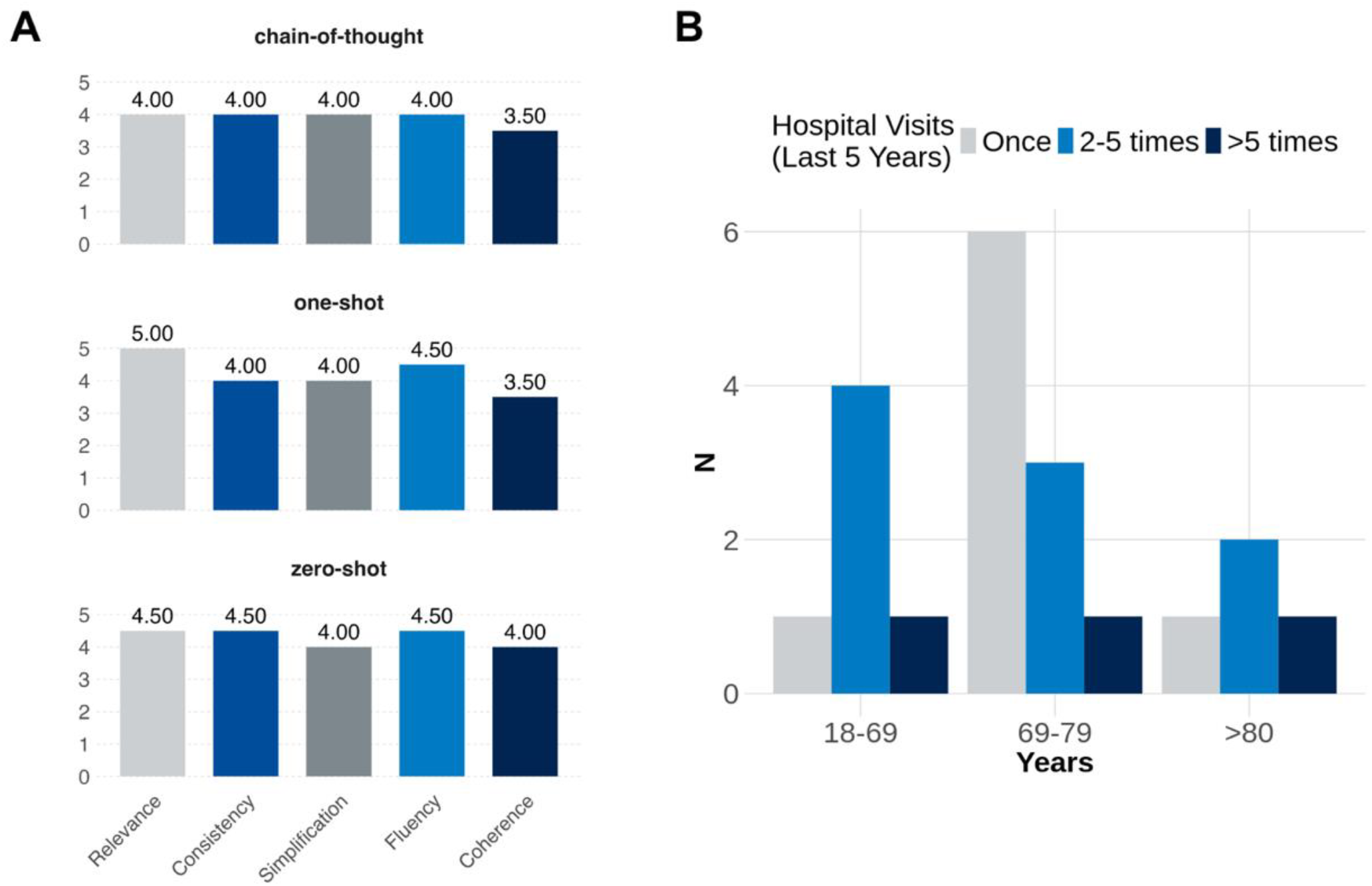
Evaluation of AI-generated content and participant characteristics. A) Human expert evaluation of outputs generated using three different prompting methods: chain-of-thought, one-shot, and zero-shot. The texts were rated on a 5-point scale across five metrics: Relevance, Consistency, Simplification, Fluency, and Coherence. The numbers above the bars indicate the mean scores. B) Distribution of the study population by age group and self-reported frequency of hospital visits within the last five years. N represents the number of participants.

### Study population

We recruited 20 participants from surgery and internal medicine departments who varied in age and medical diagnoses (Figure 1B). Half of the participants (50%, n=10/20) were aged between 69 and 79 years. A group of younger participants aged 18 to 69 years accounted for 30% (n=6/20), while the remaining 20% (n=4/20) were over 80 years old. Hospitalization frequency in the past five years (‘Once’, ‘2–5 times’, ‘>5 times’) varied across age groups (Figure 1B). Most participants (85%, n=17/20) reported either one or 2–5 hospitalizations, with only a few (15%, n=3/20) experiencing more than five. There was no association between age groups and hospitalization frequency (Chi-Square, p = 0.46).

### Patients’ baseline health literacy

Most patients reported a good understanding of the reason for their hospitalization (75%, 15/20) selecting either agree or strongly agree (Figure 2A). Three participants (15%) chose “Somewhat agree,” while two (10%) indicated “Strongly disagree”. Regarding the patients’ understanding of the examinations performed during the hospitalization, about half (55%, 11/20) agreed, seven out of 20 (35%) chose “Somewhat agree.” One participant each selected “Disagree” and “Somewhat disagree” (5% each). A similar pattern emerged for the patients’ comprehension of the therapies received and planned: eleven participants (55%) “Agreed,” one (5%) “Strongly agreed,” and five (25%) “Somewhat agreed”. Additionally, one participant each selected “Disagree”, “Strongly disagree”, and “Somewhat disagree” (5% each).

**Figure 2.**
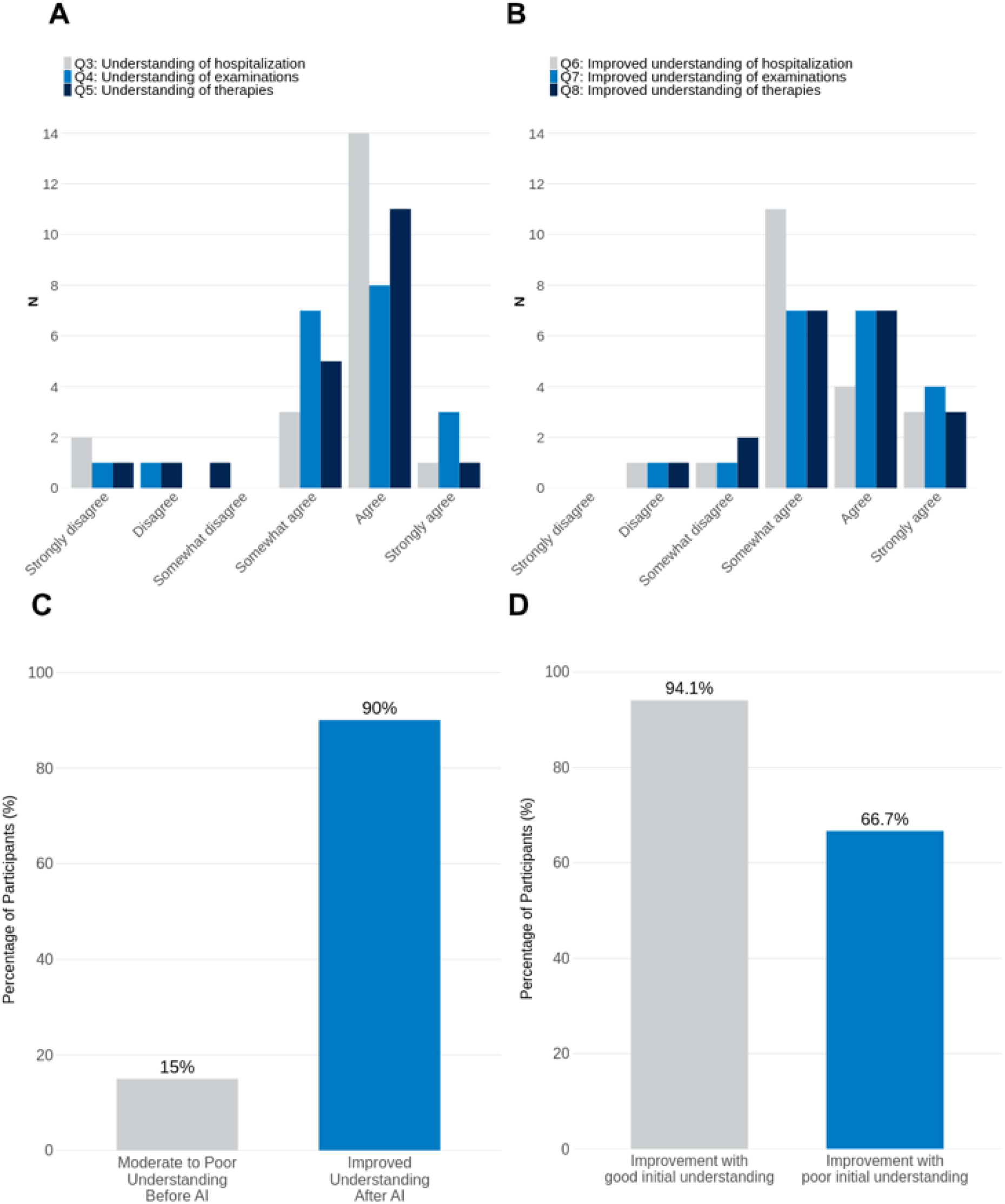
Patient understanding before and after AI intervention. A) Participants’ baseline self-assessed health literacy and B) their reported improvement after receiving AI-generated summaries. C) Percentage of participants with poor initial understanding compared to those reporting improvement after the intervention. D) Improvement rates stratified by whether the participants’ initial understanding was good or poor. In panels A/B, N represents the number of participants. AI = artificial intelligence.

The overall average understanding across all three questions (Q3, Q4, Q5) prior to reading the AI-generated summary was 4.43 out of 6 (SD = 1.21). There was no effect of the number of hospital visits (ANOVA, p = 0.426) or age (p = 0.694) on health literacy. We also excluded an interaction between age group and hospital visits (p = 0.982).

### AI-generated patient-friendly discharge summaries improve patients’ health literacy

After reading the AI-generated summary, participants were asked to assess whether their understanding had improved in the same three areas: reason for hospitalization (Q6), examinations (Q7), and therapies (Q8) (Figure 2B). Most patients reported an improved understanding of their hospitalization (90% agreed, 18/20), their examinations (90%, 18/20) and therapies (85%, 17/20).

The overall average rating for understanding following the AI-generated summary (across questions Q6, Q7, Q8) was 4.47 out of 6 (SD = 0.91). When stratified by age, this average understanding was 4.33 (SD = 0.76) for the 18-69 age group, 4.43 (SD = 1.05) for the 69-79 age group, and 4.75 (SD = 0.88) for the >80 age group. Stratifying by the number of hospital visits in the past five years, the average improvement of comprehension was 4.46 (SD = 0.50) for those visiting once, 4.30 (SD = 1.25) for those visiting 2-5 times, and 5.00 (SD = 0.33) for those visiting more than 5 times. There were no differences in these understanding scores across the different age groups (ANOVA, p = 0.785), across different frequencies of hospital visits (p = 0.533), nor was there a interaction effect between age group and hospital visits (p = 0.936).

We found that while only a few patients (15%, 3/20) had “Moderate to Poor Understanding Before AI,” the majority (90%, 18/20) of patients reported their comprehension of their medical condition improved upon reading the AI-generated summary (Figure 2C). Among those with an initial self-reported “poor” understanding, 66.7% reported improvement. Among patients with an initial self-reported “good” understanding, the majority (94.1%, 16/17) still reported an improvement, suggesting that the AI-generated summary was beneficial across varying levels of baseline comprehension (Figure 2D).

### Patient’s perception of the use of and trust in artificial intelligence in healthcare

We found that patient perspectives on AI in medicine are generally positive (Figure 3). When asked to compare the AI-generated summary with their doctor’s consultation (Q9), 90% (18/20) of participants reported that the AI-generated summary was more helpful for their comprehension. Looking towards future healthcare interactions, 85% (17/20) of patients expressed a desire to receive an AI-generated patient summary after subsequent hospital stays (Q10). Similarly, 85% of participants indicated a mainly positive attitude towards the broader application of AI in the healthcare sector (Q11). These findings underscore a high level of patient acceptance and perceived benefit from the AI-generated summaries.

**Figure 3.**
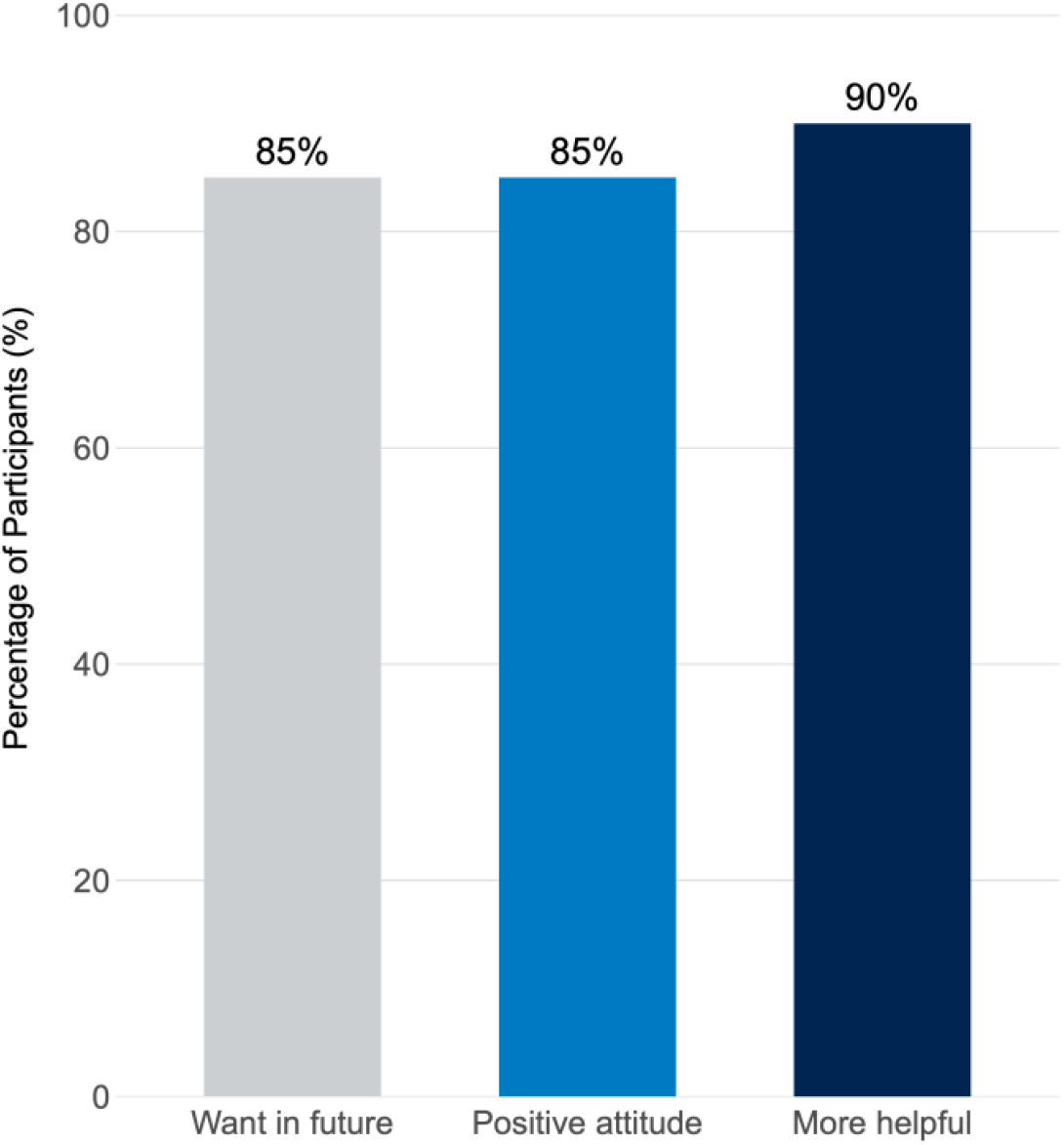
Perspectives on AI in medicine. Showing the percentage of participants who want to receive an AI-generated summary for future hospital stays, have a generally positive attitude towards AI in healthcare and found the AI-generated summary more helpful than their doctor’s discharge consultation.

While all groups reported improved comprehension with AI-generated summaries compared to physician discharge interviews and agreeing on broader use of AI in healthcare, more of the older patients indicated that they wished to receive AI-generated summaries in the future (ANOVA, p = 0.021).

## Discussion

This study demonstrated that AI-generated patient-friendly discharge summaries resulted in a self-reported improvement in health literacy for the majority of hospitalized patients. This underscores the value of AI in augmenting patient-provider communication and empowering patients in their healthcare journey.

AI-generated summaries led to a clear positive trend in patient comprehension, directly addressing our central research question and aligning with emerging research on the utility of AI in patient education (15–17). Most participants perceived their comprehension to have improved after reading the AI-generated summary, and found the summary more helpful for enhancing understanding compared to a consultation with their doctor. Notably, even patients who already rated their understanding as high prior to reading the summary still reported further improvement, highlighting the usefulness of AI-generated summaries across all levels of baseline comprehension. For all of the items, we did not find an association between age and number of hospital stays and health comprehension, suggesting that patients might benefit from AI-generated summaries independent of their health system exposure or digital literacy. This is supported by our finding that the desire to receive AI-generated summaries for future hospital stays was higher among older patients, suggesting these tools may be especially welcomed by populations often perceived as less technologically proficient. These findings support the hypothesis that AI-generated summaries enhance patient understanding and education, in line with a growing amount of evidence highlighting the benefit of AI from patients’ perspective (18,19).

One strength of this pilot study lies in its patient-centered focus, directly assessing patient-reported understanding of their medical situation, an aspect often overlooked in favor of model-centric metrics such as perplexity or clinician-verified accuracy. Another notable aspect was the practical and physician-centered methodology employed to generate the AI-generated summaries. The effectiveness of the zero-shot prompt was notable in our preliminary prompt engineering phase. This may be attributed to the inherent simplicity of zero-shot prompting, which provides the LLM with direct instructions without the potential constraints or biases introduced by few-shot examples, especially if those examples themselves contain subtle flaws or stylistic idiosyncrasies not generalizable across different medical cases (20,21). OpenAI’s GPT-4o was selected due to its widespread adoption and demonstrated high effectiveness in summarizing medical text (22).

Our study has several limitations. The small sample size (n=20) from a single academic medical center limits the generalizability of the findings. There was no control group receiving only standard means of information. The survey relied on self-reported understanding that can be influenced by politeness bias or a over- or underestimation by patients of their actual level of comprehension, as we did not objectively assess their actual health literacy / comprehension. Whether this improved understanding persists over time and translates into better long-term health behaviors is unknown.

Taken together, we provide evidence that AI-generated patient-friendly discharge summaries can enhance hospitalized patients’ self-reported health literacy. Future research should prioritize larger randomized controlled trials with objective comprehension measures, explore diverse patient populations and linguistic contexts, and systematically analyze AI accuracy and patient trust. In conclusion, AI-generated summaries demonstrate considerable promise as a powerful tool to augment clinician communication, empowering patients and potentially improving their engagement with post-discharge care.

## Data Availability

The datasets used and/or analyzed during the current study are available from the corresponding author on reasonable request.

## Declarations

### Ethics approval and consent to participate

This study was approved by the ethics committee at Charité – Universitätsmedizin Berlin (EA2/322/23).

### Competing interests

The authors declare that they have no competing interests.

### Funding

Not applicable

### Authors’ contributions

NR and VL conceived the study and performed patient recruitment, data analysis and wrote the manuscript. JJ, MW, TS, and LES oversaw patient recruitment and edited the manuscript. MG supervised the study and edited the manuscript.

## Notes

### Competing Interest Statement

The authors have declared no competing interest.

### Funding Statement

This study did not receive any funding

### Author Declarations

This study was approved by the ethics committee at Charite Universitaetsmedizin Berlin (EA2/322/23).

## References

1. Kessels RPC. Patients’ memory for medical information. J R Soc Med. 2003 May;96(5):219–22.

2. Jansen J, Butow PN, van Weert JCM, van Dulmen S, Devine RJ, Heeren TJ, et al. Does age really matter? Recall of information presented to newly referred patients with cancer. J Clin Oncol. 2008 Nov 20;26(33):5450–7.

3. Warner JL, Levy MA, Neuss MN, Warner JL, Levy MA, Neuss MN. ReCAP: Feasibility and accuracy of extracting cancer stage information from narrative electronic health record data. J Oncol Pract. 2016 Feb;12(2):157–8; e169-7.

4. Hvalvik S, Dale B. The transition from hospital to home: Older people’s experiences. Open J Nurs. 2015 Jul 9;05(07):622–31.

5. Holstead RG. Utility of large language models to produce a patient-friendly summary from oncology consultations. JCO Oncol Pract. 2024 Sep;20(9):1157–9.

6. Abreu AA, Murimwa GZ, Farah E, Stewart JW, Zhang L, Rodriguez J, et al. Enhancing readability of online patient-facing content: The role of AI chatbots in improving cancer information accessibility. J Natl Compr Canc Netw. 2024 May 15;22(2 D):1–8.

7. Van Veen D, Van Uden C, Blankemeier L, Delbrouck J-B, Aali A, Bluethgen C, et al. Adapted large language models can outperform medical experts in clinical text summarization. Nat Med. 2024 Apr;30(4):1134–42.

8. Aydin S, Karabacak M, Vlachos V, Margetis K. Large language models in patient education: a scoping review of applications in medicine. Front Med (Lausanne). 2024 Oct 29;11:1477898.

9. Pristoupil J, Oleaga L, Junquero V, Merino C, Sureyya OS, Kyncl M, et al. Generative pre-trained transformer 4o (GPT-4o) in solving text-based multiple response questions for European Diploma in Radiology (EDiR): a comparative study with radiologists. Insights Imaging. 2025 Mar 22;16(1):66.

10. Best practices for prompt engineering with the OpenAI API [Internet]. [cited 2025 May 19]. Available from: https://help.openai.com/en/articles/6654000-best-practices-for-prompt-engineering-with-the-openai-api

11. R: A language and environment for statistical computing. R Foundation for Statistical Computing. Vienna, Austria; 2023.

12. Wickham H, Averick M, Bryan J, Chang W, McGowan L, François R, et al. Welcome to the tidyverse. J Open Source Softw. 2019 Nov 21;4(43):1686.

13. likert: Analysis and Visualization Likert Items [Internet]. Comprehensive R Archive Network (CRAN). [cited 2025 Jun 12]. Available from: https://CRAN.R-project.org/package=likert

14. Patil I. Visualizations with statistical details: The “ggstatsplot” approach. J Open Source Softw. 2021 May 25;6(61):3167.

15. Stanceski K, Zhong S, Zhang X, Khadra S, Tracy M, Koria L, et al. The quality and safety of using generative AI to produce patient-centred discharge instructions. NPJ Digit Med. 2024 Nov 20;7(1):329.

16. Conrado F, Rosset L, Lo Moro G, Scaioli G, Consoli D, Bert F, et al. Effect on comprehension of an AI patient-friendly hospital discharge letter: a quasi-RCT. Eur J Public Health [Internet]. 2024 Nov 1;34(Supplement_3). Available from: 10.1093/eurpub/ckae144.685

17. De Rouck R, Wille E, Gilbert A, Vermeersch N. Assessing artificial intelligence-generated patient discharge information for the emergency department: a pilot study. Int J Emerg Med. 2025 Apr 25;18(1):85.

18. Ayers JW, Poliak A, Dredze M, Leas EC, Zhu Z, Kelley JB, et al. Comparing physician and artificial intelligence chatbot responses to patient questions posted to a public social media forum. JAMA Intern Med. 2023 Jun 1;183(6):589–96.

19. Chen S, Guevara M, Moningi S, Hoebers F, Elhalawani H, Kann BH, et al. The effect of using a large language model to respond to patient messages. Lancet Digit Health. 2024 Jun;6(6):e379–81.

20. Sivarajkumar S, Kelley M, Samolyk-Mazzanti A, Visweswaran S, Wang Y. An empirical evaluation of prompting strategies for large language models in zero-shot clinical natural language processing: Algorithm development and validation study. JMIR Med Inform. 2024 Apr 8;12:e55318.

21. Kojima T, Gu SS, Reid M, Matsuo Y, Iwasawa Y. Large Language Models are Zero-Shot Reasoners [Internet]. arXiv [cs.CL]. 2022. Available from: http://arxiv.org/abs/2205.11916

22. Liu C-L, Ho C-T, Wu T-C. Custom GPTs enhancing performance and evidence compared with GPT-3.5, GPT-4, and GPT-4o? A study on the Emergency Medicine Specialist Examination. Healthcare [Internet]. 2024 Aug 30;12. Available from: 10.3390/healthcare12171726

