## Supplementary File S1 for "AI-generated patient-friendly discharge summaries to empower patients"

### Prompt Evaluation

Name: ______________________________ Date: ____________________

The evaluations are based on a 5-point Likert scale, where 1 stands for "very poor" and 5 stands for "very good." Detailed explanations of the categories can be found in the attached letter.

| **Case 1.1** | 1 | 2 | 3 | 4 | 5 |
|---|---|---|---|---|---|
| Relevance | ☐ | ☐ | ☐ | ☐ | ☐ |
| Consistency | ☐ | ☐ | ☐ | ☐ | ☐ |
| Simplification | ☐ | ☐ | ☐ | ☐ | ☐ |
| Fluency | ☐ | ☐ | ☐ | ☐ | ☐ |
| Coherence | ☐ | ☐ | ☐ | ☐ | ☐ |

Notes:

| **Case 1.2** | 1 | 2 | 3 | 4 | 5 |
|---|---|---|---|---|---|
| Relevance | ☐ | ☐ | ☐ | ☐ | ☐ |
| Consistency | ☐ | ☐ | ☐ | ☐ | ☐ |
| Simplification | ☐ | ☐ | ☐ | ☐ | ☐ |
| Fluency | ☐ | ☐ | ☐ | ☐ | ☐ |
| Coherence | ☐ | ☐ | ☐ | ☐ | ☐ |

Notes:
