## Supplementary File S2 for "AI-generated patient-friendly discharge summaries to empower patients"

### Questionnaire on Experiences with Artificial Intelligence in Healthcare

**Please check the applicable items.**

1. **I belong to the following age group:**
   - ☐ 18-49 years
   - ☐ 50-69 years
   - ☐ 69-79 years
   - ☐ >80 years

2. **How often have you been hospitalized in the past 5 years?**
   - ☐ Once
   - ☐ 2-5 times
   - ☐ >5 times

Please read the following statements carefully and assess how much you agree with them.

3. **After the conversation with my attending physician, I have a good understanding of the reason for my hospitalization.**
   - ☐ Strongly disagree
   - ☐ Disagree
   - ☐ Somewhat disagree
   - ☐ Somewhat agree
   - ☐ Agree
   - ☐ Strongly agree

4. **After the conversation with my attending physician, I have a good understanding of the examinations carried out during my hospital stay and why they were necessary.**
   - ☐ Strongly disagree
   - ☐ Disagree
   - ☐ Somewhat disagree
   - ☐ Somewhat agree
   - ☐ Agree
   - ☐ Strongly agree

5. **After the conversation with my attending physician, I have a good understanding of the therapies administered and planned during my hospital stay and why they are necessary.**
   - ☐ Strongly disagree
   - ☐ Disagree
   - ☐ Somewhat disagree
   - ☐ Somewhat agree
   - ☐ Agree
   - ☐ Strongly agree

Please now read the AI-generated patient summary. Then read the following statements and assess how much you agree with them.

6. **After reading the AI-generated patient summary, I understand the reason for my hospitalization better than before.**
   - ☐ Strongly disagree
   - ☐ Disagree
   - ☐ Somewhat disagree
   - ☐ Somewhat agree
   - ☐ Agree
   - ☐ Strongly agree

7. **After reading the AI-generated patient summary, I understand the examinations carried out and the reason they were necessary better than before.**
   - ☐ Strongly disagree
   - ☐ Disagree
   - ☐ Somewhat disagree
   - ☐ Somewhat agree
   - ☐ Agree
   - ☐ Strongly agree

8. **After reading the AI-generated patient summary, I understand the therapies administered and planned for my condition better than before.**
   - ☐ Strongly disagree
   - ☐ Disagree
   - ☐ Somewhat disagree
   - ☐ Somewhat agree
   - ☐ Agree
   - ☐ Strongly agree

9. **Compared to the doctor's conversation, the AI summary helped me understand better.**
   - ☐ Strongly disagree
   - ☐ Disagree
   - ☐ Somewhat disagree
   - ☐ Somewhat agree
   - ☐ Agree
   - ☐ Strongly agree

10. **In the future, I would also like to receive an AI-generated patient summary after my hospital stay.**
   - ☐ Strongly disagree
   - ☐ Disagree
   - ☐ Somewhat disagree
   - ☐ Somewhat agree
   - ☐ Agree
   - ☐ Strongly agree

11. **I have a positive attitude towards the use of artificial intelligence in healthcare.**
   - ☐ Strongly disagree
   - ☐ Somewhat disagree
   - ☐ Neither agree nor disagree
   - ☐ Somewhat agree
   - ☐ Strongly agree
